# Impact of *APOE* genotype on cognition in idiopathic and genetic forms of Parkinson’s disease

**DOI:** 10.1101/2022.11.06.22281991

**Authors:** Christos Koros, Kathrin Brockmann, Athina-Maria Simitsi, Anastasia Bougea, Hui Liu, Ann-Kathrin Hauser, Claudia Schulte, Stefanie Lerche, Ioanna Pachi, Nikolaos Papagiannakis, Roubina Antonelou, Athina Zahou, Isabel Wurster, Efthymia Efthymiopoulou, Ion Beratis, Matina Maniati, Marina Moraitou, Helen Michelakakis, Georgios Paraskevas, Sokratis G. Papageorgiou, Constantin Potagas, Dimitra Papadimitriou, Maria Bozi, Maria Stamelou, Thomas Gasser, Leonidas Stefanis

## Abstract

**Background:** Apolipoprotein E-ε4 (*APOE* ε4) genotype may be associated with the development of cognitive decline in idiopathic Parkinson’s disease i(PD), however its effect in genetic PD is understudied.

**Objectives:** In the current work we aimed to assess the impact of *APOE* genotype on cognition in iPD as well as in genetic PD with mutations in the *Alpha-synuclein* (*SNCA*) and *Glycocerebrosidase* (*GBA1*) genes.

**Methods:** Two independent PD cohorts were analyzed: The first cohort (Athens) included 50 iPD patients, 35 patients with the p.A53T *SNCA* mutation and 59 patients with *GBA1* mutations (13 mild /46 severe). The second cohort (Tübingen) included 292 patients with *GBA1* mutations (170 risk/ 52 mild/ 70 severe). All patients underwent cognitive testing and were genotyped for *APOE*.

**Results:** In the iPD subgroup, carriers of at least one *APOE* ε4 exhibited lower Montreal Cognitive Assessment test (MoCA) score as compared to non-carriers (p=0.044). Notably, in the p.A53T *SNCA* subgroup, *APOE* ε4 carriers also had lower MoCA scores compared to non-carriers (p=0.039). There were no *APOE* ε4-related differences in the two *GBA1* subgroups (Athens, p=0.729; Tübingen p=0.585).

**Conclusions:** We confirm the impact of *APOE* ε4 on cognitive decline in iPD and for the first time report a similar effect in p.A53T *SNCA* mutation carriers, who represent the prototypical genetic synucleinopathy. Contrary, the lack of such an effect in two independent cohorts of *GBA1* mutation carriers, who are thought to also manifest a predominant alpha-synuclein-driven cognitive decline, suggests differences in factors associated with cognitive dysfunction between different genetic forms of synucleinopathies.

## 1. Introduction

The ε4 allele of the apolipoprotein (*APOE*) gene is the most common genetic risk factor for late onset sporadic Alzheimer’s disease (AD). The apolipoprotein E (APOE) protein, coded by the *APOE* gene, is involved in lipid transport, metabolism, and inflammatory modulation ^1^. The frequencies of the *APOE* alleles (*APOE* ε2, *APOE* ε3 and *APOE* ε4) in the general population vary slightly between different populations, however the *APOE* ε3 is the most abundant and the *APOE* ε2 is the least common ^2^. In Europe, there is a gradient of *APOE* ε4 allele distribution among populations, with high ε4 allele prevalence in northern Europe (approximately 25%) and low allele prevalence in the Mediterranean area (less than 10%) ^3^. While the *APOE* ε4 allele is a major risk factor for AD, the *APOE* ε2 allele appears to have a protective role. The AD risk is dose dependent. From a molecular perspective, *APOE* is implicated in mechanisms related to amyloid beta (Aβ) aggregation and impaired clearance in the brain. Literature evidence supports an association between *APOE* ε4 and brain Aβ burden. In particular, *APOE* ε4 was reported to directly promote Aβ fibrillization, but it may also contribute to AD pathogenesis via Aβ-independent mechanisms ^2^. *APOE* ε4 interacts with tau pathology in AD as homozygous *APOE* ε4 carriers have higher tau pathology compared to heterozygotes or non-carriers. APOE fragments which originate from *APOE* ε4 specifically can directly affect mitochondrial function ^1^. Finally, there is also evidence that neuroinflammation, having a key role in the pathogenesis of many neurodegenerative diseases, may influence the crosstalk between *APOE* ε4 and amyloid beta or tau protein.

More recently, *APOE* is also increasingly associated with the development of cognitive decline in idiopathic Parkinson’s disease (PD). It has been proposed that *APOE* ε4 increases the risk and lowers the age of onset for motor manifestation of PD as well as for PD dementia ^4-7^ but its role in PD is still debated as other studies found no firm associations between *APOE* and PD features ^8,9^. Moreover, the *APOE* locus is strongly associated with risk for developing other synucleinopathies such as dementia with Lewy bodies (DLB) [10]. In a study assessing the impact of *APOE* polymorphisms and mutations in the *Glucocerebrosidase* gene (*GBA1*) on DLB in an Ashkenazi Jewish population, it appeared that both factors had distinct effects on the clinical disease phenotype, separately and in combination ^11^.

Cognitive dysfunction in genetic forms of PD is variable, depending on the affected gene, the particular mutation, but also on other factors such as additional genetic modifiers, lifestyle, co-morbidities and environmental components. There is marked heterogeneity in the expressivity of the dementia phenotype between different genetic forms of PD and even among members of the same family ^12^. Patients harboring mutations in the genes *Alpha-synuclein* (*SNCA*) and *GBA1* often have a more pronounced cognitive burden than those with mutations in *Parkin (PRKN) or Leucine-rich repeat kinase 2 (LRRK2)* ^12,13^. The putative role of *APOE* as a genetic modifier of cognitive trajectories in genetic forms of PD is largely unexplored. The aim of our current study was to assess the impact of *APOE* genotype on cognitive function in iPD patients and in patients with pathogenic mutations in the genes *SNCA* and *GBA1* as these two PD subgroups show a prominent and early cognitive decline during the disease course ^14,15^.

## 2. Materials and Methods

We have analyzed two independent PD cohorts. The first cohort included patients followed in the Movement Disorders Outpatient Clinic of Eginition Hospital, University of Athens (Greece). We assessed 50 idiopathic PD patients, 35 patients with the p.A53T *SNCA* mutation and 59 patients with *GBA1* mutations (13 mild /46 severe according to mutation severity as classified for risk of PD). The second cohort included 292 patients with *GBA1* mutations (170 risk/ 52 mild/ 70 severe) followed in the Outpatient Clinic for Parkinson’s Disease at the University of Tübingen (Germany). The present study was conducted in agreement with the principles of the Declaration of Helsinki. Written informed consent was obtained from all participants. Raw data used in the current study are available upon request.

Epidemiological data including PD age at onset (defined as manifestation of motor onset) were collected (Table 1). All patients were genotyped for *APOE* at the University of Tübingen. Moreover, patients underwent cognitive testing using the Montreal Cognitive Assessment test (MoCA) ^16^ [a subset had undergone testing with the Mini Mental State Examination Test (MMSE) which was then converted to MoCA according to published algorithms ^17^].

**Table 1.** Demographic data and MoCA score of the 3 Athens cohort subgroups

| | Idiopathic<br>PD_no $\epsilon 4$<br>n=44 | Idiopathic<br>PD_at least one $\epsilon 4$<br>n=6 | p.A53T<br>SNCA_no $\epsilon 4$<br>n=29 | p.A53T<br>SNCA_at least one<br>$\epsilon 4$<br>n=6 | GBA1_no $\epsilon 4$<br>n=49 | GBA1_at<br>least one $\epsilon 4$<br>n=10 |
| --- | --- | --- | --- | --- | --- | --- |
| Male sex, % (n) | 66(29) | 33(2) | 45(13) | 50(3) | 53(26) | 60(6) |
| Age | 66.34±1.31 | 64.67±3.28 | 50.8±2.12 | 57±3.34 | 59.7±1.81 | 56.6±2.49 |
| Age at onset | 59.25±1.45 | 59.33±3.57 | 44.66±2.2 | 50±3.2 | 51.1±1.59 | 50±2.88 |
| Disease duration | 7.43±0.85 | 5.33±0.85 | 6.14±0.8 | 7±1.87 | 8.91±0.81 | 6.6±1.63 |
| Education | 12.83±0.6 | 12.67±2.19 | 13.92±0.78 | 8.67±1.15 | 10.88±0.71 | 13±1.34 |
| MoCA score | 24.71± 0.55 | 22.17± 1.94 | 24.41±0.95 | 16.17±3 | 23.1±0.93 | 23.2±1.82 |

MoCA: Montreal Cognitive Assessment test, SNCA: α-synuclein gene, GBA1: Glucocerebrosidase gene

Additionally, patients from the Tübingen cohort have also undergone analysis of CSF biomarkers [including amyloid beta (Abeta1_42), total Tau protein (t-Tau), phosphorylated Tau protein (p181-Tau), neurofilament light chain (NFL) and α-synuclein].

Statistical analysis was performed using one-way ANCOVA for continuous variables (with age, sex, disease duration, education as covariates) and Pearson chi-square for categorical variables. Statistical significance was set at p<0.05.

## 3. Results

The frequency of presence of at least one *APOE*ε4 allele (either in heterozygous or in homozygous form) was 6/50 (12%) in the idiopathic PD subgroup and 6/35 (17.14%) in the p.A53T *SNCA* PD subgroup. The frequency of *APOE*ε4 allele in the Athens *GBA1* PD cohort was 10/59 (16.95%) and in the Tübingen *GBA1* PD cohort 72/292 (24.66%) This difference is consistent to the latitude gradient effect of *APOE* ε4 frequency, with lower frequencies observed in Mediterranean countries as opposed to Northern Europe countries ^3^ (Figure 1).

**Figure 1.**
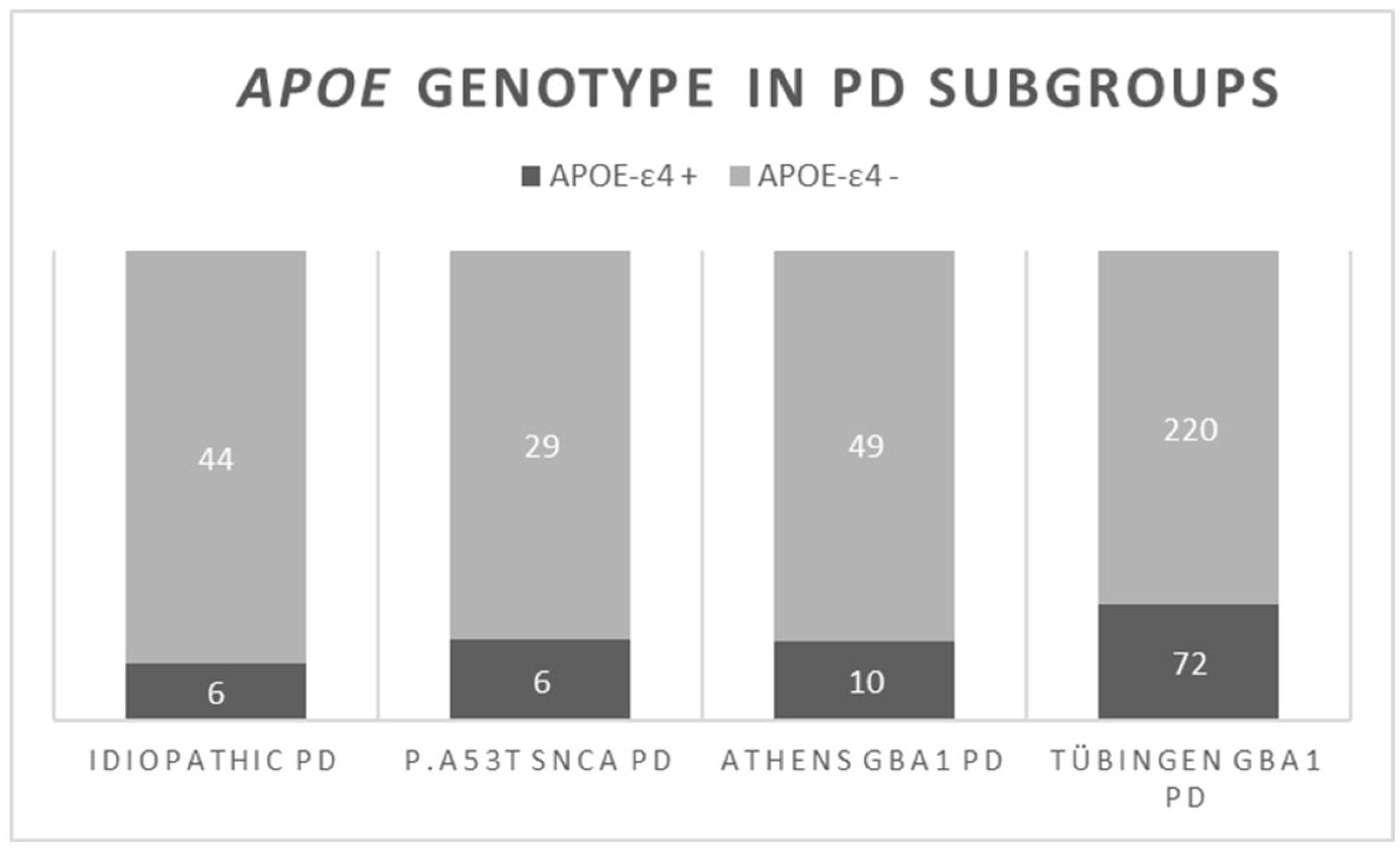
The frequency of presence of at least one *APOE* ε4 allele (either in heterozygous or in homozygous form) in the idiopathic PD subgroup and in the genetic subgroups (p.A53T *SNCA* PD and *GBA1* PD)

The cognitive status of PD patients as exemplified by their MoCA score presented a marked variability in each subgroup: Demographic data and MoCA scores are provided in Table 1 and Table 2.

**Table 2:**
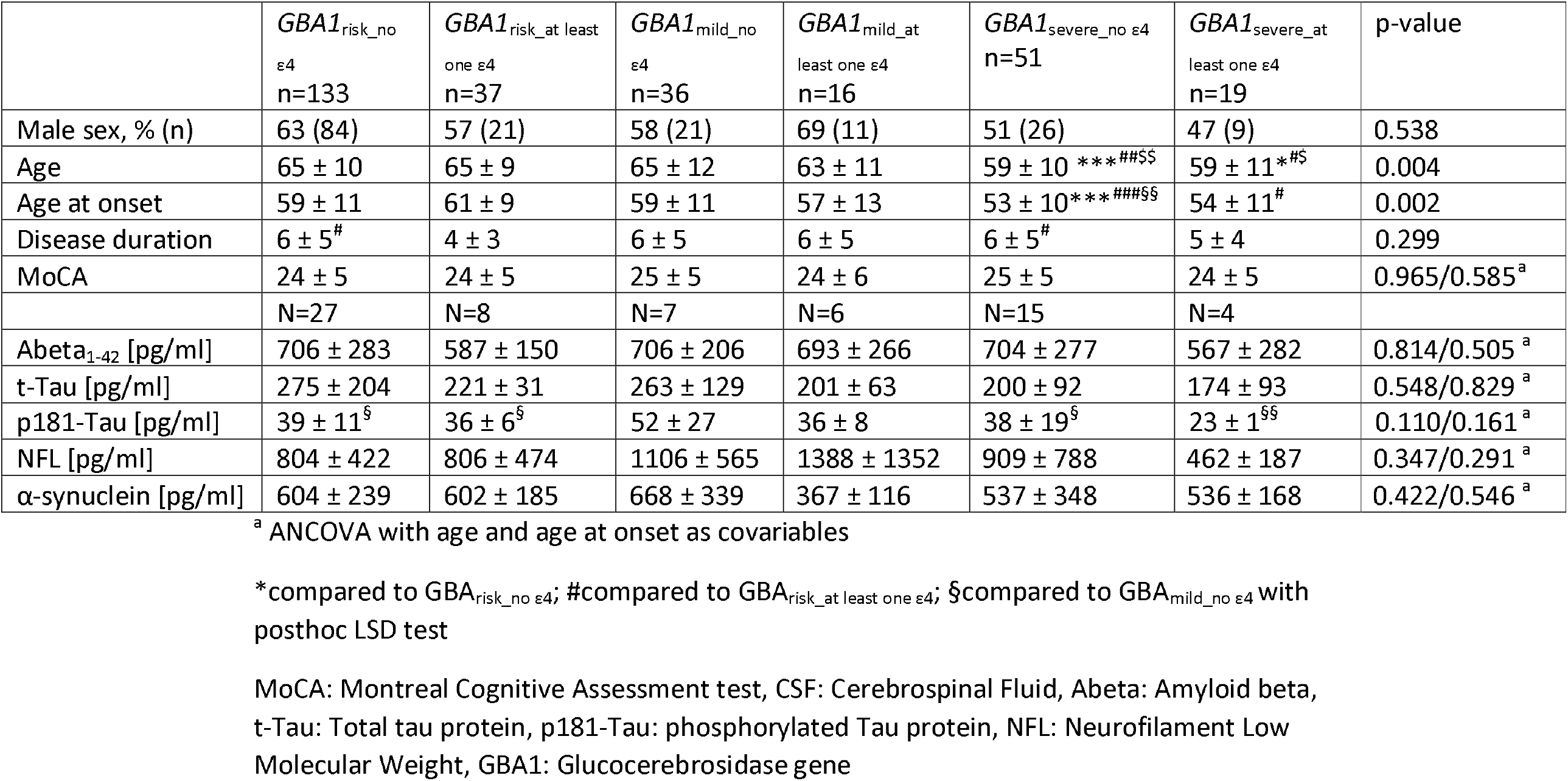
Demographic, clinical and CSF Biomarker in Tübingen PD_GBA_ (first visit with MoCA)

In the iPD subgroup of the Athens cohort, carriers of at least one *APOE* ε4 exhibited lower MoCA score as compared to non-carriers when using age, sex, disease duration and education as covariates (22.2 vs 24.7/30) (p=0.044). Moreover, no difference was noticed regarding the age at PD onset [*APOE*ε4+ (59.33) vs *APOE*ε4-(59.25) p= 0.501 with age and sex as covariates] (Figure 2A).

**Figure 2.**
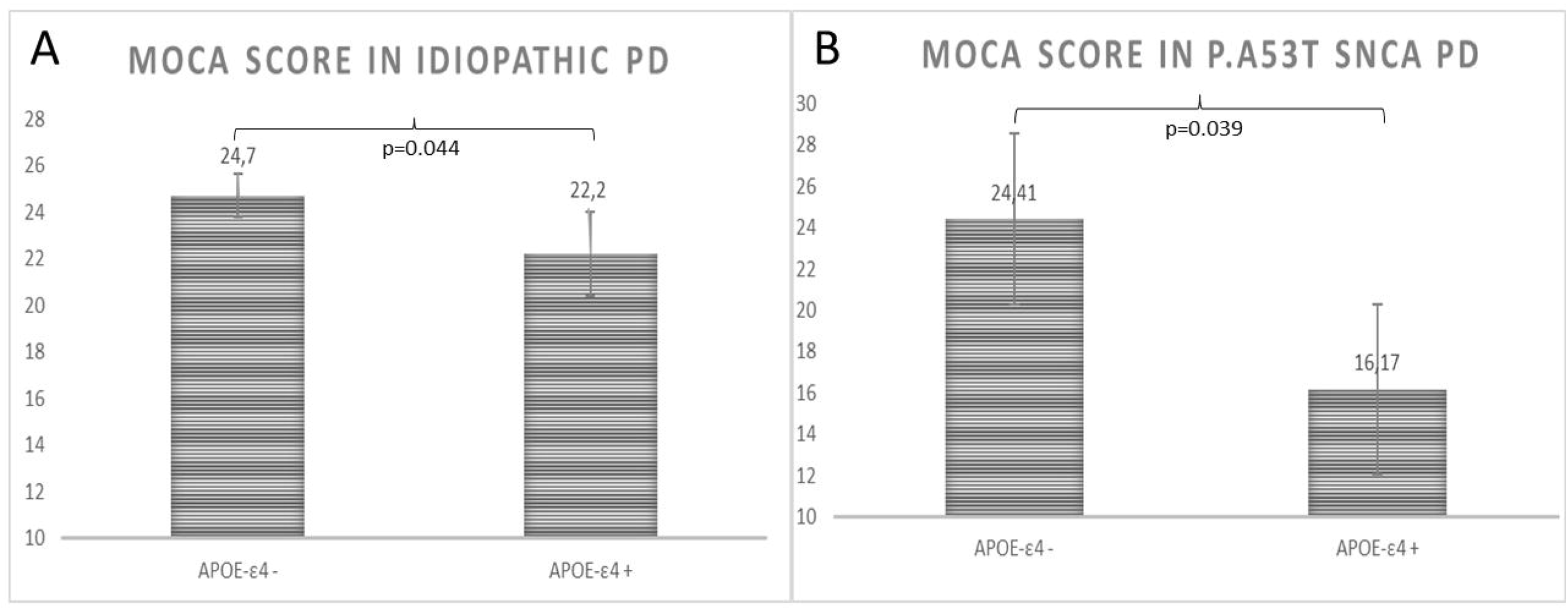
MoCA scores in *APOE* ε4 carriers and non-carriers of the Idiopathic PD subgroup (Fig 2A). MoCA scores in *APOE* ε4 carriers and non-carriers of the p.A53T *SNCA* PD subgroup (Fig 2B).

Notably, in the p.A53T *SNCA* subgroup, *APOE* ε4 carriers also had lower MoCA scores compared to non-carriers (16.2 vs 24.4/30) (p=0.039). Furthermore, no difference was noticed regarding the age at PD onset [*APOE* ε4+ (50) vs *APOE* ε4-(44.7) p= 0.736 with age and sex as covariates] (Figure 2B).

When assessing cognitive status as a categorical variable (cognitive decline corresponding to MoCA cutoff <26/30), the difference was significant only for the p.A53T *SNCA* (p=0.014) but not for iPD (p=0.443).

In the *GBA1* subgroup from the Athens cohort, no difference in MoCA score related to *APOE* ε4 status was detected (p=0.647). This was also true when the severity of *GBA1* mutations was used as covariate (p=0.729). No difference was noticed regarding the age at PD onset [*APOE* ε4+ (50) vs *APOE* ε4-(51.2) p=0.227 with age, sex and mutation severity as covariates] (Figure 3A).

**Figure 3.**
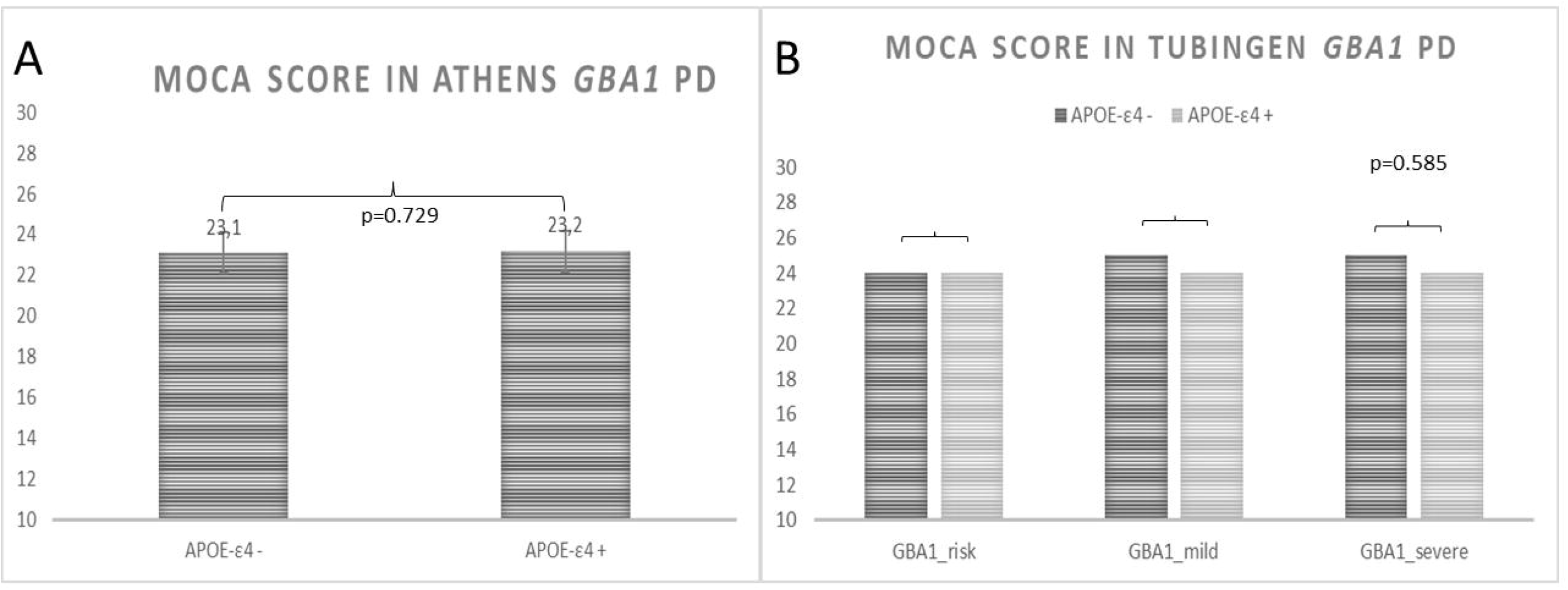
MoCA scores in *APOE* ε4 carriers and non-carriers of the Athens *GBA1* PD subgroup (Fig 3A). MoCA scores in *APOE* ε4 carriers and non-carriers of the Tubingen *GBA1* PD subgroup (Fig 3B).

Accordingly, in the Tübingen *GBA1* PD cohort there was no difference between *APOE*ε4 carriers and non-carriers when assessing the three subgroups of *GBA1* variant severity (risk, mild, severe p=0.585) (Figure 3B) (Table 2)

Moreover, there were no associations between *APOE* genotype and CSF measurement outcomes [total amyloid beta Abeta1-42 (p= 0.505), t-Tau (p=0.829), p181-Tau (p=0.161), NFL (p=0.291), α-synuclein (p=0.546)] with age and age at onset as covariables (Table 2 and Supplementary Table 3).

## 4. Discussion

In this cross-sectional study we found a negative effect of *APOE* ε4 allele on the global cognitive function of idiopathic PD patients as exemplified by the MoCA test scores. Epidemiological and educational factors have been taken into account in order to avoid any possible bias. Our results are in accordance with literature data regarding the already described impact of *APOE* ε4 on cognitive decline of iPD. Common genetic variation of the *APOE* and other genes including micro-tubule associated protein tau (*MAPT*) have been linked to cognitive decline and dementia in Parkinson’s disease, although former studies have yielded mixed results. A study evaluating 152 neuropathologically confirmed Parkinson’s disease donors with or without clinical dementia during life concluded that both the *APOE* ε4 allele and *MAPT* H1-haplotype were associated with earlier development of dementia ^18^. Male sex and *APOE* ε4, along with age and lower education level, were associated with poorer cognitive performance [using Mattis Dementia Rating Scale (DRS-2)] among a population of predominantly non-demented PD patients ^19^. A magnetic resonance imaging study showed that the Functional Connectivity (FC) values between the bilateral caudate and posterior cortical regions of *APOE* ε4 carriers with PD-MCI were much lower than *APOE* ε3 and ε2 allele carriers, and FC was positively correlated with the impairment of global and language function ^20^. A very recent study focused on DLB and might shed additional light on the *APOE* effect. Kaivola and co-authors performed a comprehensive evaluation in 2466 dementia with LB cases using an APOE-stratified genome-wide association study approach. They concluded that *APOE* ε4 effect was associated with mixed dementia due to mixed LB/ AD pathology but not with pure dementia with LB ^10^.

The *APOE* ε4 effect was even more prominent in longitudinal studies. A 7-year observational study from Norway showed that PD patients carriers of the *APOE* ε4 allele had an increased risk of mild cognitive impairment and dementia within the study period ^21^. According to results from the Parkinson’s Progression Markers Initiative (PPMI) study, *APOE* status along with baseline age, University of Pennsylvania Smell Inventory Test (UPSIT) scores and CSF amyloid beta - (Aβ42) to t-tau ratio were associated with change in MoCA scores over time in de novo PD patients ^22^. Data from de novo PD patients indicate that CSF Aβ-42 and *APOE* ε4 promote early cognitive changes in PD patients as those with low CSF Aβ-42 and *APOE* ε4+ showed a higher rate of cognitive decline early in the disease course ^23^. Finally, in a sporadic PD cohort from Tuebingen, low levels of CSF amyloid beta were associated with a higher risk of developing cognitive impairment earlier in the disease course ^6,7^.

In our current report, the *APOE* ε4 effect on cognitive dysfunction was also present in the p.A53T *SNCA* subgroup, which represents the prototypical genetic synucleinopathy. *APOE* ε4 carriers in this genetic cohort exhibited significantly lower MoCA scores and were more often characterized as demented, based on MOCA cut-offs, compared to non-carriers. The degree of cognitive deterioration in PD patients with the p.A53T *SNCA* mutation ranges from intact cognition to severe Parkinson’s Disease Dementia (PDD). Regarding the pattern of cognitive decline, frontal-executive and visuospatial functions are more prominently affected ^24^. The idea that *APOE* could act as an additional genetic modifier of cognitive impairment in this genetic *SNCA* cohort is thus plausible. Despite literature evidence indicating that *APOE* ε4 directly exacerbates α-synuclein pathological deposition, beyond its established role in promoting AD co-pathology, we were unable to provide CSF biomarker data (amyloid beta and alpha-synuclein) regarding our *SNCA* cohort. Previous data from our group, have shown low amyloid beta in a proportion of *p*.*A53T SNCA* PD carriers ^25^.

The *APOE* ε4 genotype mechanism of action in cognitive impairment in PD is not clear, because patients often have a mixture of α-synuclein (αSyn), amyloid beta (Aβ), and tau pathologies. *APOE* ε4 exacerbates brain Aβ pathology, as well as tau pathology, but it is not clear whether *APOE* genotype independently regulates alpha-synuclein pathology ^2^. A recent study using a mouse model of alpha-synucleinopathy by means of adeno-associated virus gene delivery of α-synuclein in human APOE-targeted replacement mice expressing *APOE* ε2, *APOE* ε3, or *APOE* ε4 showed that only APOE ε4 increased α-synuclein pathology and impaired behavioral performances. The results revealed a pathogenic role of *APOE* ε4 in exacerbating α-synuclein pathology independent of amyloid beta ^26^. Furthermore, a second research group generated A53T *SNCA* transgenic mice on *APOE* knockout or human *APOE* knockin backgrounds and demonstrated that that *APOE* genotype directly regulates αSyn pathology independent of its established effects on Aβ and tau ^27^. Finally, another study has found indications of ApoE fragmentation in the human PD brain and documented the presence of APOE within Lewy bodies, the major PD pathological marker ^28^. From a clinical perspective, a study group assessed autopsy confirmed Lewy Body brains in order to decipher whether *APOE* ε4 is associated with severity of LB pathology, independently of AD pathology. They reached the conclusion that *APOE* ε4 is independently associated with a greater severity of LB pathology and that *APOE* ε4 may function as a modifier of processes that favor LB spread rather than acting directly to initiate LB pathology ^29^.

Regarding the *GBA*1 subgroups, cognitive impairment is more frequent and severe in *GBA1* PD as compared to iPD and the cognitive decline appears earlier in the disease course. However, a marked variability in cognitive impairment is also encountered among different *GBA1* mutations ^30^. Interestingly, we did not find an effect of *APOE* genotype on cognitive function in two independent cohorts of *GBA1* mutation carriers, who are considered a model of alpha-synuclein-related cognitive decline. The absence of differences in CSF biomarkers Abeta1_42, t-Tau, p181-Tau and alpha-synuclein profiles between *APOE* ε4 carriers and non-carriers in the *GBA1* cohort further supports that the *APOE* genotype is irrelevant in this genetic PD form.

The joint results from our current study and former research suggest differences in factors associated with cognitive decline (genetic, epigenetic or environmental) between different genetic forms of synucleinopathies, even those that would be expected to be closely related, such as is the case of *GBA1* and *SNCA* mutations. In this respect, carriers of the p.A53T *SNCA* mutation resemble more closely iPD compared to *GBA1* mutation carriers, given the presence of low CSF beta-amyloid levels in some cases ^25^ and the association of cognitive decline with the *APOE* ε4 genotype reported here.

## Data Availability

All data produced in the present study are available upon reasonable request to the authors

## Table captions

Supplementary Table 3. Demographic, clinical and CSF Biomarker in Tübingen *GBA1* PD (first visit with CSF collection and biomarker measurement)

## Authors’ Roles

Christos Koros, Research project: A. Conception, B. Organization, C. Execution; Statistical Analysis: C. Review and Critique; Manuscript: A. Writing of the first draft, B. Review and Critique.

Kathrin Brockmann, Research project: A. Conception, B. Organization, C. Execution; Statistical Analysis: C. Review and Critique; Manuscript: A. Writing of the first draft, B. Review and Critique.

Athina Simitsi, Research project: B. Organization, C. Execution; Statistical Analysis: A. Design, B. Execution; Manuscript: B. Review and Critique.

Anastasia Bougea, Research project: B. Organization, C. Execution; Statistical Analysis: A. Design, B. Execution; Manuscript: B. Review and Critique.

Hui Liu, Research project: B. Organization, C. Execution; Statistical Analysis: C. Review and Critique; Manuscript: B. Review and Critique.

Ann-Kathrin Hauser, Research project: B. Organization, C. Execution; Statistical Analysis: C. Review and Critique; Manuscript: B. Review and Critique.

Claudia Schulte, Research project: B. Organization, C. Execution; Statistical Analysis: C. Review and Critique; Manuscript: B. Review and Critique.

Stefanie Lerche, Research project: B. Organization, C. Execution; Statistical Analysis: C. Review and Critique; Manuscript: B. Review and Critique.

Ioanna Pachi, Research project: B. Organization, C. Execution; Statistical Analysis: C. Review and Critique; Manuscript: B. Review and Critique.

Nikolaos Papagiannakis, Research project: B. Organization, C. Execution; Statistical Analysis: C. Review and Critique; Manuscript: B. Review and Critique.

Roubina Antonelou, Research project: B. Organization, C. Execution; Statistical Analysis: C. Review and Critique; Manuscript: B. Review and Critique.

Athina Zahou, Research project: B. Organization, C. Execution; Statistical Analysis: C. Review and Critique; Manuscript: B. Review and Critique.

Isabel Wurster, Research project: B. Organization, C. Execution; Statistical Analysis: C. Review and Critique; Manuscript: B. Review and Critique.

Efthymia Efthymiopoulou, Research project: B. Organization, C. Execution; Statistical Analysis: C. Review and Critique; Manuscript: B. Review and Critique.

Ion Beratis, Research project: Research project: B. Organization, C. Execution; Statistical Analysis: C. Review and Critique; Manuscript: B. Review and Critique.

Matina Maniati, Research project: B. Organization, C. Execution; Statistical Analysis: C. Review and Critique; Manuscript: B. Review and Critique.

Marina Moraitou, Research project: B. Organization, C. Execution; Statistical Analysis: C. Review and Critique; Manuscript: B. Review and Critique.

Helen Michelakakis, Research project: B. Organization, C. Execution; Statistical Analysis: C. Review and Critique; Manuscript: B. Review and Critique.

Georgios Paraskevas, Research project: B. Organization, C. Execution; Statistical Analysis: C. Review and Critique; Manuscript: B. Review and Critique.

Sokratis G. Papageorgiou, Research project: B. Organization; Statistical Analysis: C. Review and Critique; Manuscript: B. Review and Critique.

Dimitra Papadimitriou, Research project: B. Organization, C. Execution; Statistical Analysis: C. Review and Critique; Manuscript: B. Review and Critique.

Maria Bozi, Research project: Research project: B. Organization, C. Execution; Statistical Analysis: C. Review and Critique; Manuscript: B. Review and Critique.

Maria Stamelou, Research project: B. Organization, C. Execution; Statistical Analysis: C. Review and Critique; Manuscript: B. Review and Critique.

Thomas Gasser, Research project: A. Conception, B. Organization, C. Execution; Statistical Analysis: C. Review and Critique; Manuscript: B. Review and Critique.

Leonidas Stefanis, Research project: A. Conception, B. Organization, C. Execution; Statistical Analysis: C. Review and Critique; Manuscript: A. Writing of the first draft, B. Review and Critique.

Financial Disclosures of all authors (for the preceding 12 months)

Christos Koros received funding from the Michael J Fox Foundation for his participation in PPMI.

Kathrin Brockmann is employed at the University of Tuebingen and served as Consultancy and Advisory Board in Hoffmann La Roche. She has received honoraria as a Speaker from UCB and ABBVie and a Grant from the MJFF.

Athina Simitsi received funding from the Michael J Fox Foundation for her participation in PPMI and from “ALAMEDA” study (H2020-EU, Grant Agreement 101017558.

Anastasia Bougea received funding from “ALAMEDA” study (H2020-EU, Grant Agreement 101017558.

Hui Liu has no disclosures.

Ann-Kathrin Hauser has no disclosures.

Claudia Schulte has no disclosures.

Stefanie Lerche has no disclosures.

Ioanna Pachi has no disclosures.

Nikolaos Papagiannakis has no disclosures.

Roubina Antonelou has no disclosures.

Athina Zahou has no disclosures.

Isabel Wurster has no disclosures.

Efthymia Efthymiopoulou received funding from the Michael J Fox Foundation for her participation in PPMI.

Ion Beratis received funding from the Michael J Fox Foundation for his participation in PPMI.

Matina Maniati has no disclosures.

Marina Moraitou has no disclosures.

Helen Michelakakis has no disclosures.

Georgios Paraskevas has no disclosures.

Sokratis G. Papageorgiou has no disclosures.

Constantin Potagas has no disclosures

Dimitra Papadimitriou has no disclosures. Maria Bozi has no disclosures.

Maria Stamelou, serves on the editorial boards of Movement Disorders Journal and Frontiers in Movement Disorders and has received research support from the Michael J Fox Foundation (PPMI).

Thomas Gasser has received a grant from Bundesministerium für Bildung und Forschung (BMBF) Heinemannstrasse 2 und 6, 53170 Bonn. Moreover he has received funding from Deutsche Forschungsgemeinschaft (DFG), from the Ministry for Science, Research and Art Baden-Württemberg (MWK) and from The Michael J. Fox Foundation for Parkinson’s Research (Steering Committee Member Award). Prof. Gasser has received speakers honoraria from UCB Pharma, Novartis, Teva and MedUpdate. He is chairman of the scientific advisory board of the “Joint Programming for Neurodegenerative Diseases” program, funded by the European Commission.

Leonidas Stefanis over the past year has received the following grants : PPMI2 (supported by the Michael J. Fox Foundation), IMPRIND-IMI2 Number 116060 (EU, H2020), “Transferring autonomous and non-autonomous cell degeneration 3D models between EU and USA for development of effective therapies for neurodegenerative diseases (ND) - CROSS NEUROD” (H2020-EU 1.3.3., Grant Number778003), «Chaperone-Mediated Autophagy in Neurodegeneration» (Hellenic Foundation for Research and Innovation Grant HFRI-FM17-3013), “ALAMEDA” (H2020-EU, Grant Agreement 101017558), “Next-generation antisense molecules for Parkinson’s disease therapy (THERASYN)” (Greek Secretariat of Research and Technology (GSRT), Collaborator), “National Network for Neurodegenerative Diseases (EΔIAN)” (GSRT), and “CMA as a Means to Counteract alpha-Synuclein Pathology in Non-Human Primates” (by the Michael J. Fox Foundation (Collaborator)). He has served on Advisory Boards for Abbvie, ITF Hellas and Biogen and has received honoraria from Abbvie and Sanofi. There are no specific disclosures related to the current work.

## Notes

Financial Disclosure/Conflict of Interest Christos Koros received funding from the Michael J Fox Foundation for his participation in PPMI.

### Competing Interest Statement

The authors have declared no competing interest.

### Funding Statement

This study was partially funded by the National Network for Neurodegenerative Diseases (ΕΔΙΑΝ) (GSRT).

### Author Declarations

Eginition Hospital Ethics Committee Ethical Aproval was granted by the EC for the conduction of the study as a part of Thalis Parkinson's disease database

