## Supplementary Table 3 for "Impact of *APOE* genotype on cognition in idiopathic and genetic forms of Parkinson’s disease"

Supplementary Table 3: Demographic, clinical and CSF Biomarker in PD_GBA_ (first visit with Biomarker)

|  | GBA_risk_no ε4_  n=47 | GBA_risk_at least one ε4_  n=12 | GBA_mild_no ε4_  n=16 | GBA_mild_at least one ε4_  n=10 | GBA_severe_no ε4_  n=21 | GBA_severe_at least one ε4_  n=10 | p-value |
| --- | --- | --- | --- | --- | --- | --- | --- |
| Male sex, % (n) | 68 (32) | 67 (8) | 63 (10) | 70 (7) | 67 (14) | 60 (6) | 0.995 |
| Age | 67 ± 9 | 64 ± 7 | 69 ± 11 | 62 ± 10 | 58 ± 10 ***^$$$^ | 60 ± 9^$^ | 0.004 |
| Age at onset | 59 ± 10 | 59 ± 7 | 60 ± 10 | 55 ± 11 | 50 ± 11***^#§§^ | 52 ± 11 | 0.010 |
| Disease duration | 8 ± 5^#^ | 4 ± 3 | 9 ± 5^#^ | 8 ± 5 | 8 ± 6^#^ | 8 ± 6 | 0.259 |
| MoCA | 23 ± 5 | 24 ± 6 | 24 ± 6 | 26 ± 4 | 25 ± 5 | 24 ± 6 | 0.867/0.587^a^ |
| Abeta_1-42_ [pg/ml] | 754 ± 272^#^ | 552 ± 147 | 657 ± 218 | 645 ± 266 | 721 ± 257 | 651 ± 279 | 0.210/0.252 ^a^ |
| t-Tau [pg/ml] | 263 ± 162 | 253 ± 143 | 276 ± 157 | 204 ± 64 | 198 ± 90 | 177 ± 74 | 0.232/0.729 ^a^ |
| p181-Tau [pg/ml] | 40 ± 12^§^ | 41 ± 23 | 51 ± 24 | 36 ± 11^§^ | 37 ± 17^§§^ | 28 ± 10^§§^ | 0.034/0.195 ^a^ |
| NFL [pg/ml] | 874 ± 514 | 883 ± 506 | 872 ± 462 | 1287 ± 1226 | 805 ± 691 | 942 ± 553 | 0.570/0.373 ^a^ |
| α-synuclein [pg/ml] | 588 ± 222 | 576 ± 172 | 651 ± 290 | 360 ± 120*^§§^ | 520 ± 325 | 560 ± 113 | 0.153/0.254 ^a^ |

^a^ ANCOVA with age and age at onset as covariables

*compared to GBA_risk_no ε4_; #compared to GBA_risk_at least one ε4_; §compared to GBA_mild_no ε4_ with posthoc LSD test

MoCA: Montreal Cognitive Assessment test, Abeta: Amyloid beta, t-Tau: Total tau protein, p181-Tau: phosphorylated Tau protein, NFL: Neurofilament Low Molecular Weight
